# Using Whole Genome Sequences to Investigate Adenovirus Outbreaks, Including Five Deaths in a Haematopoietic Stem Cell Transplant Unit

**DOI:** 10.1101/2020.11.26.20239111

**Authors:** Chloe E. Myers, Charlotte J. Houldcroft, Sunando Roy, Ben K. Margetts, Timothy Best, Cristina Venturini, José A. Guerra-Assunção, Charlotte A. Williams, Rachel Williams, Helen Dunn, John C. Hartley, Kathryn J. Rolfe, Judith Breuer

**Affiliations:** Cambridge Clinical Microbiology and Public Health Laboratory, Public Health England, Cambridge, United Kingdom; Department of Medicine, University of Cambridge, Cambridge, United Kingdom; Division of Infection and Immunity, University College London, United Kingdom; Division of Infection, Immunity and Inflammation, University College London, Great Ormond Street Institute of Child Health, United Kingdom; Great Ormond Street Hospital for Children NHS Foundation Trust, London, United Kingdom

**Keywords:** adenovirus, epidemiology, whole-genome sequencing, paediatric infectious disease, nosocomial transmission

## Abstract

**Background:** Human mastadenoviruses (HAdV) are associated with significant morbidity and mortality amongst the immunocompromised population. A recent surge in HAdV cases, including five deaths, amongst a haematopoietic stem cell transplant population led us to use whole genome sequencing (WGS) to investigate.

**Methods:** To gain a complete transmission picture, we compared outbreak and non-outbreak sequences (54 sequences from 37 patients) with GenBank sequences and our own database of previously sequenced HAdVs (132 sequences from 37 patients). An improved bait set for WGS was used. Maximum likelihood trees and pairwise differences were used to evaluate genotypic relationships paired with epidemiological data from routine Infection, Prevention and Control (IPC) activity.

**Results:** Nine monophyletic clusters were identified, seven of which were corroborated by epidemiological evidence and by comparison of single nucleotide polymorphisms. Two incomplete patient clusters were identified by IPC over the same time period. Of the five patients who died, one had a mixed HAdV infection and two were the source of transmission events.

**Conclusions:** The clinical consequences of unmitigated HAdV transmission events are high. Focusing on two high risk wards using WGS we identified six transmission events, over prolonged periods, that would have gone unnoticed using traditional polymerase chain reaction and epidemiology. Mixed infection is frequent (10% of patients), providing a sentinel source of recombination and superinfection. Immunosuppressed patients harbouring a high rate of HAdV positivity require comprehensive surveillance. As a consequence of these findings, HAdV WGS is being incorporated routinely into a clinical algorithm to prevent transmission and influence IPC policy in real-time.

**Summary:** Whole genome sequencing of adenovirus, direct from clinical samples, can be used to identify cryptic health care associated transmission events, and to resolve transmission suspected by traditional epidemiology. It can also identify mixed genotype infections in immunocompromised patient populations.

## Background

Clinical infections caused by human mastadenoviruses (HAdVs) are associated with significant morbidity (10-89%) and mortality (6-70%) in the immunocompromised host (1). Risk factors for poor outcome include paediatric patients (who are susceptible to primary infection), unrelated donor stem cell transplants (SCTs), graft-versus host disease, T-cell depletion of graft and certain immunosuppressive drug regimens (2–4).

The burden of HAdV infection is significant; within the paediatric oncology population HAdV has been reported to account for 15% of all diarrhoeal cases (5). Amongst paediatric patients undergoing haematopoietic stem cell transplant (HSCT), HAdV viraemia and stool shedding was found in 15% and 42% of patients respectively (6,7). As non-enveloped viruses, HAdVs can be resistant to standard alcohol cleaning regimens and can survive as clinically infectious particles for up to four weeks (8). Nosocomial transmission has been frequently reported in the literature (9,10) however, identification of these outbreaks is likely to be under-reported due to limitations of existing HAdV typing protocols that are performed infrequently and target only small regions of selected genes (11).

Advances in whole genome sequencing (WGS) have provided valuable insights into the molecular epidemiology of a number of key hospital pathogens (12–15). This has been well illustrated recently in the context of severe acute respiratory syndrome coronavirus 2 (SARS-CoV-2), where application in real-time has allowed prompt feedback supporting epidemiological links and the utility of existing IPC policies (16).

Specifically within our population, a tertiary paediatric referral centre of which 30% of patients are immunocompromised, HAdV is one of the leading causes of viral gastroenteritis, comprising 44% of all infections (17). Over the last financial year (2019-2020) there were 642 new HAdV detections, from any sample site, 99 of which were new viraemias (local audit data). Adenoviraemia significantly decreases the probability of survival in children following HSCT and also increases the duration of inpatient hospital stay, with an associated financial burden (6,18–20). Due to the volume of HadV infections there is often no clear indication whether infection is acquired from the community, hospital, or reactivation from a latent site, making it very difficult to interrupt spread.

In our hospital over a 20-month period, seven HAdV outbreaks have been investigated by the IPC team; two were associated with the HSCT unit and included five deaths. This was despite rigorous IPC policies including environmental screening (21,22). In response, we undertook extensive epidemiological investigation and sequencing of isolates from the HSCT unit to determine what proportion were transmitted. Using WGS data we document the genetic relatedness between isolates and describe possible transmission events. These findings can be used to interrupt HAdV transmission dynamics, by influencing routine IPC policy and improved patient care.

## Methods

### Context and Ethics

Great Ormond Street Hospital (GOSH) is a 350 bed, paediatric tertiary referral centre. Due to the immunocompromised status of patients referred here, over 60% of beds are single room isolation facilities. In addition to those patients who are symptomatic, ‘high-risk’ patients - those admitted for haematological transplant or congenital immunodeficiencies – are screened weekly and on admission for gastrointestinal infection using polymerase chain reaction (PCR). PCR methods used by the GOSH diagnostic laboratory have been described previously (23). Residual diagnostic samples were collected from patients with PCR confirmed HAdV infection. The PCR cycle threshold (C_*T*_) values provided a comparable semiquantitative indicator of viral titre. Use of these samples for research was approved by The National Research Ethics Service Committee London – Fulham (reference: 17/LO/1530). Clinical data was extracted from hospital databases by the GOSH Digital research Environment (DRE) team and linked to an anonymised patient number.

### Definitions, Patients and Samples

A HSCT unit nosocomial outbreak is suspected when any new detection of HAdV infection is identified in a child who was negative on admission screening. Further information on the routine management of outbreaks are provided in Supplementary Methods. For surveillance reporting, healthcare acquired infection (HCAI) is defined as a positive diagnostic sample ≥48 hours post admission; and community-acquired infection (CAI), a positive diagnostic sample within 48 hours of admission and no healthcare contact in the preceding 14 days.

A total of 169 samples from 74 patients were included in the study (Supplementary Table 1). All patients were known to have either a congenital or acquired immunodeficiency and therefore considered high-risk. As part of this investigation, 11 outbreak samples (*n* = eight patients) identified as two clusters by IPC (infection control cluster one (ICC one), patients: 54, 55, 56, 57, 62, 68 and infection control cluster two (ICC two), patients: 40 and 38) and 37 non-outbreak samples (*n* = 29 patients, including HCAI and CAI infections), were sequenced and analysed with a local database of HAdV sequences (127 sequences from 37 patients).

### SureSelect Bait Design & Sequencing

Methods allowing high-throughput HAdV WGS directly from clinical samples have been developed (23–25). These methods provide a proof of concept that WGS offers the resolution required to confirm nosocomial transmission of HAdV however, there were technical improvements to be made with species C viruses (85/107 clinical samples) yielding lower quality sequences (23). 120-mer baits (version 2) were redesigned, using an in-house perl script with a tiling factor of 12x (each position in a given genome is covered by 12 unique bait designs), against all whole HAdV sequences (487) in GenBank (accessed Jan 24, 2018). The bait design was uploaded to SureDesign and biotinylated RNA oligonucleotides (baits) were synthesised by Agilent Technologies (26).

Quality control of sample DNA, library preparation using the SureSelect^XT^ Illumina paired-end protocol and sequencing on an Ilumina MiSeq sequencer were performed as described previously (23), except we utilised the SureSelect^XT^ low input kit. Base calling and sample demultiplexing were performed as standard for the MiSeq platform, generating paired FASTQ files for each sample [GenBank accessions XXXX-XXXX].

### Genome Mapping, Assembly and Phylogenetic Analysis

Sequences for all 169 samples were assembled using a reference-based pipeline in CLC Genomics Workbench version 12.0.1 (QIAGEN); detailed methodology can be found in Supplementary methods and Supplementary Figure 1. Briefly, all reads were quality trimmed and adaptor sequences removed. Trimmed reads were mapped to a reference database (n = 103), where 90% of each read mapped with a minimum of 90% identity, the best reference match was used to assign a genotype to each sample. If mapped reads generated a good match to more than one genotype, suggesting a mixed infection, samples underwent further investigation [Supplementary methods, Supplementary Figures 1-5 and Supplementary Tables 2-6].

Once a sample had been assigned a genotype a second pipeline was then used to quality trim, re-map to the best reference match with a length and similarity fraction of 0.8, before extracting a consensus sequence. Areas of low coverage (<10 fold) were assigned the ambiguity symbol N. Robust consensus sequences are required for downstream analysis, therefore only samples achieving ≥90% genome coverage and ≥100-fold average read depth (quality cut-off) were included in further analysis. Consensus sequences were aligned, and phylogenies constructed using CLC Genomics Workbench (version 12.0.1) [Supplementary methods]. Pairwise single-nucleotide variant counts were computed using Molecular Evolutions Genetics Analysis (MEGA) software version six (27).

### Epidemiological Support of Phylogenetic Clusters

Monophyletic clusters, defined as groups comprising two or more samples from at least two patients arising from a common ancestral node, with bootstrap support ≥90% were used to identify putative outbreaks. Timelines for each monophyletic cluster were visualised using the ggplot 2 library (28), incorporating patient admission data and HAdV PCR positivity. Patients within a cluster were defined as epidemiologically supported if they were present on the same ward or unit becoming positive during the incubation period of the virus (median 5.6 days (95% CI 4.8-6.3) based on respiratory disease (29)), and unsupported if they became positive during admissions to completely different wards or had no links with any other sequenced patient.

### Statistical Analysis

Statistical tests were performed using two-tailed tests at the 5% significance level within GraphPad Prism version 8.3.0 for mac OS, GraphPad Software, San Diego, California USA, www.graphpad.com [Supplementary methods].

## Results

### Burden of Infection and Viral Genotypes

Routine reporting of first PCR positive HAdV cases by the diagnostic laboratory between 2015 and 2019 is summarised in Figure 1. As expected, no seasonality was observed (17), however cases increased each year with a marked rise between 2018-2019. The proportion of new cases that were attributed to HCAI during this time period rose from 12% (2015-2017) to 23% (2018-2019) (Figure 1B). On average 18% of new positives are detected from patients admitted to the high-risk HSCT unit (Figure 1C).

Between August 2017 and April 2019 IPC identified 11 outbreak samples (from ICC one and ICC two) and 37 non-outbreak samples (including HCAI and CAI cases) from high-risk patients. Sequences were analysed with 121 previously sequenced samples. A total of 169 clinical samples containing HAdV genotypes A31 (14%), B3 (2%), B11 (1%), C1 (17%), C2 (21%), C5 (18%), C89 (8%), E4 (1%) and F41 (3%) from 74 patients with either localised (eye, respiratory, digestive) or disseminated infection were included. Seven of these samples (4%) failed to sequence and 17 (10% of patients) had mixed HAdV infections (Supplementary Table 1 and Supplementary Figure 6).

### Improved Sequencing Quality

Of 169 samples across all genotypes, 56 (42%) of 132 HAdV genomes passed the quality cut-off using version one baits and 46 (85%) of 54 HAdV genomes passed using version two baits (Supplementary Table 1 and Supplementary Figure 7). Both genome coverage and on-target reads (OTRs) were statistically significantly improved for species C viruses using version two baits [Supplementary Figure 8]. This was despite similar species C viral titres in samples between bait groups [Supplementary Figure 9]. Average read depth improved but remained significantly lower for species C viruses regardless of the baits used (*P* = 0.0002 version one baits, versus *P* = 0.05, version two baits).

Of the seven samples that failed to sequence, four were sequenced using version one baits. Three samples failed using version two baits, one with a viral load that had previously been successful. There was insufficient sample for repeat testing. Using an estimated linear regression model, it is predicted that samples with HAdV C_*T*_ values of ≤34 would generate a ≥100 fold read depth, with 95% certainty [Supplementary Figure 10].

### Phylogenetic Investigation of Outbreaks and Deaths

To substantiate nosocomial transmission, maximum likelihood phylogenies were constructed, Figure 2. Nine monophyletic clusters were identified (Figures 2A to 2E) and summarised in Table 1. One of these clusters (A31 Cluster four) had been identified phylogenetically previously (23).

**Figure 1.**
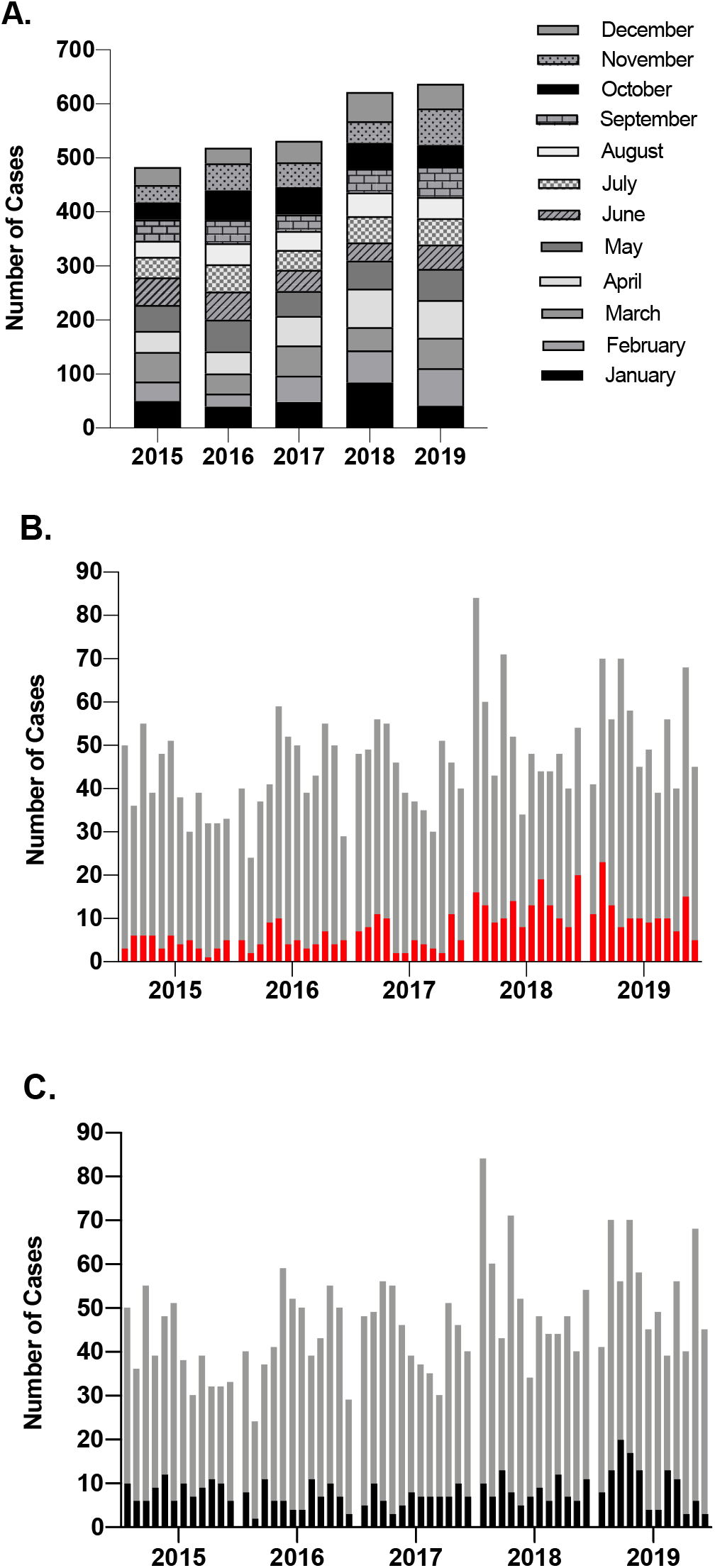
Incidence of new HAdV cases diagnosed using PCR by the diagnostic laboratory. (A.) The total number of HAdV cases identified by GOSH increased each year but did not demonstrate any seasonality. (B.) The proportion of new positives that were documented as HCAI are shown in red, and (C.) the proportion of new positives identified from patients admitted to the high-risk unit are highlighted in black. Abbreviations: HAdV, human mastadenovirus; PCR, polymerase chain reaction; GOSH, Great Ormond Street Hospital; HCAI, healthcare-acquired infection.

**Figure 2.**
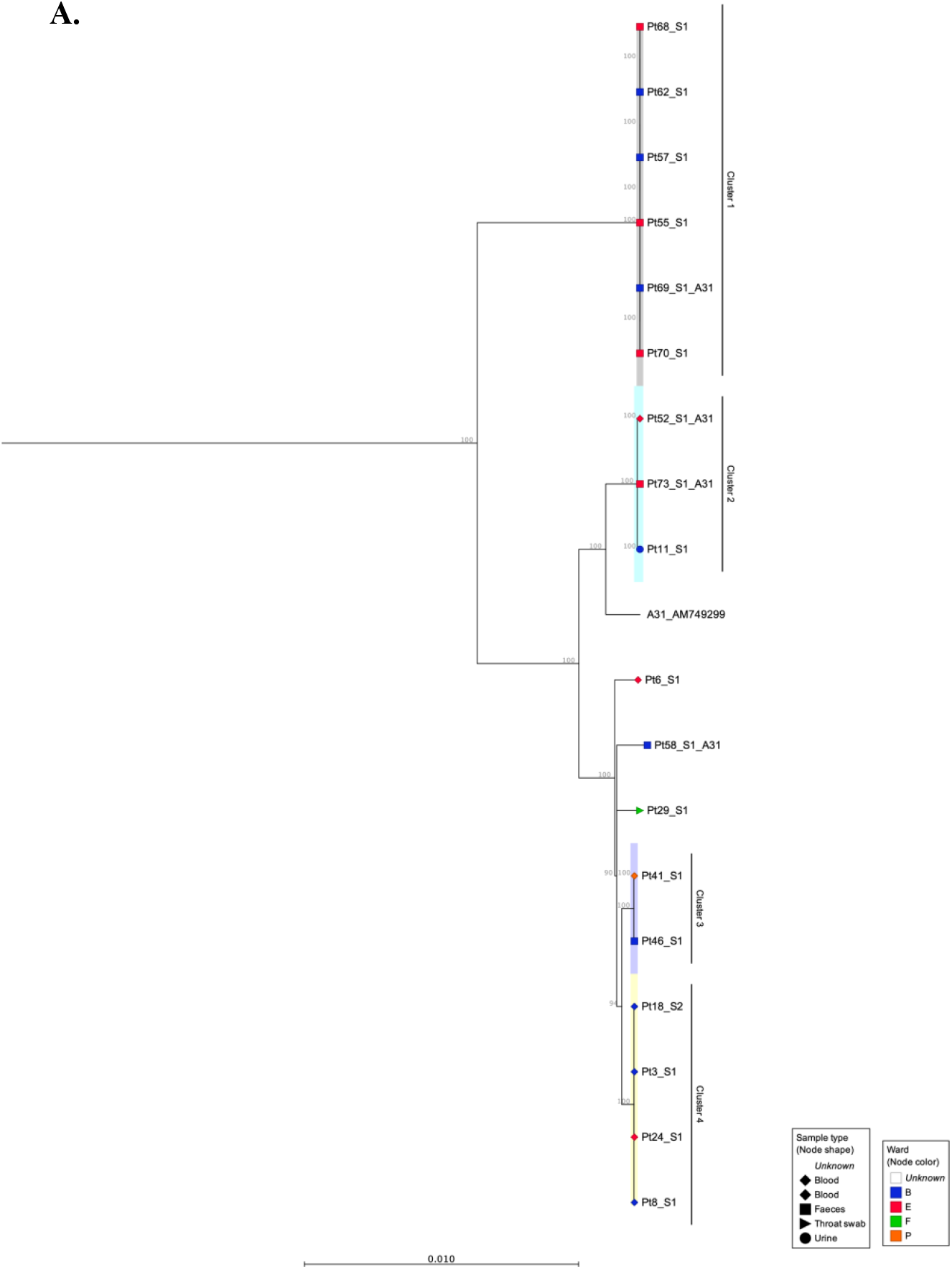
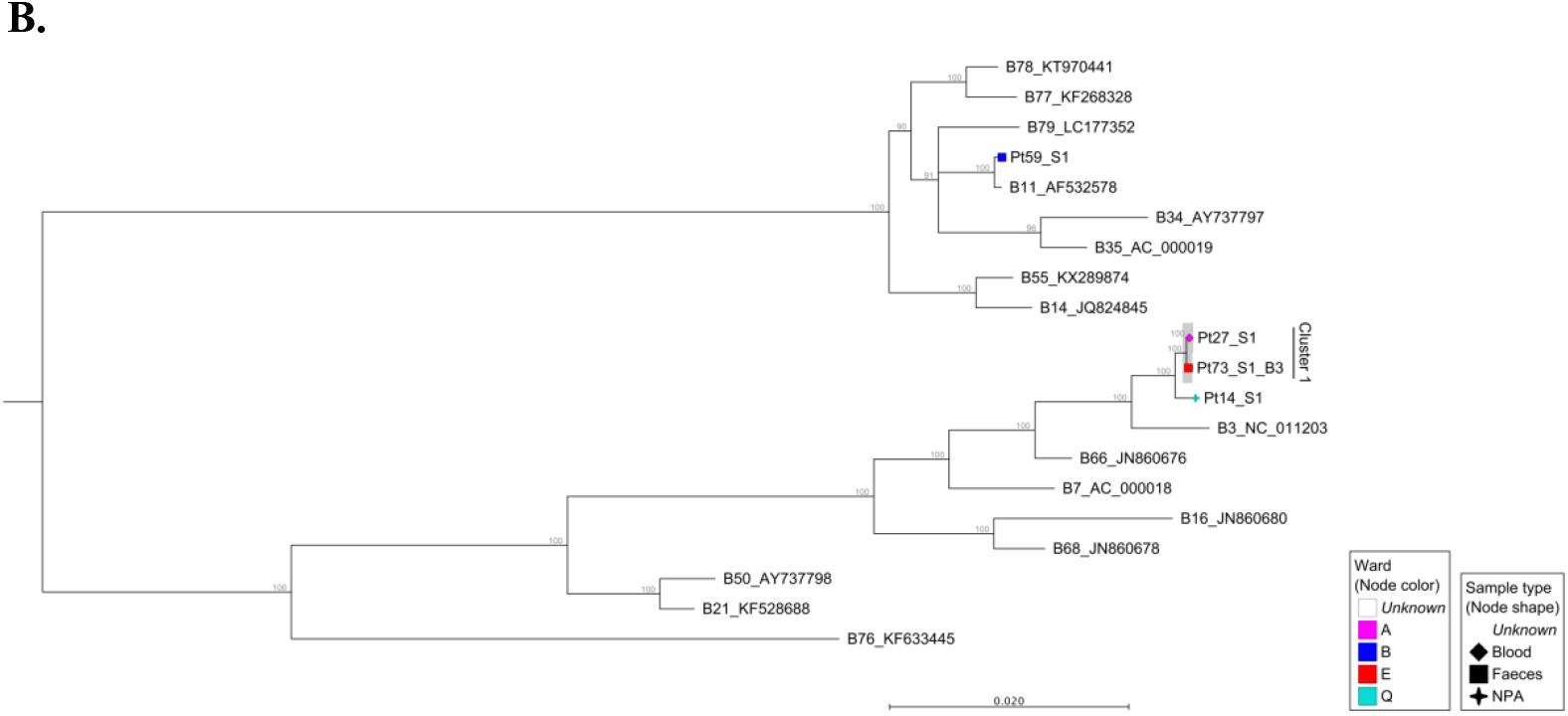
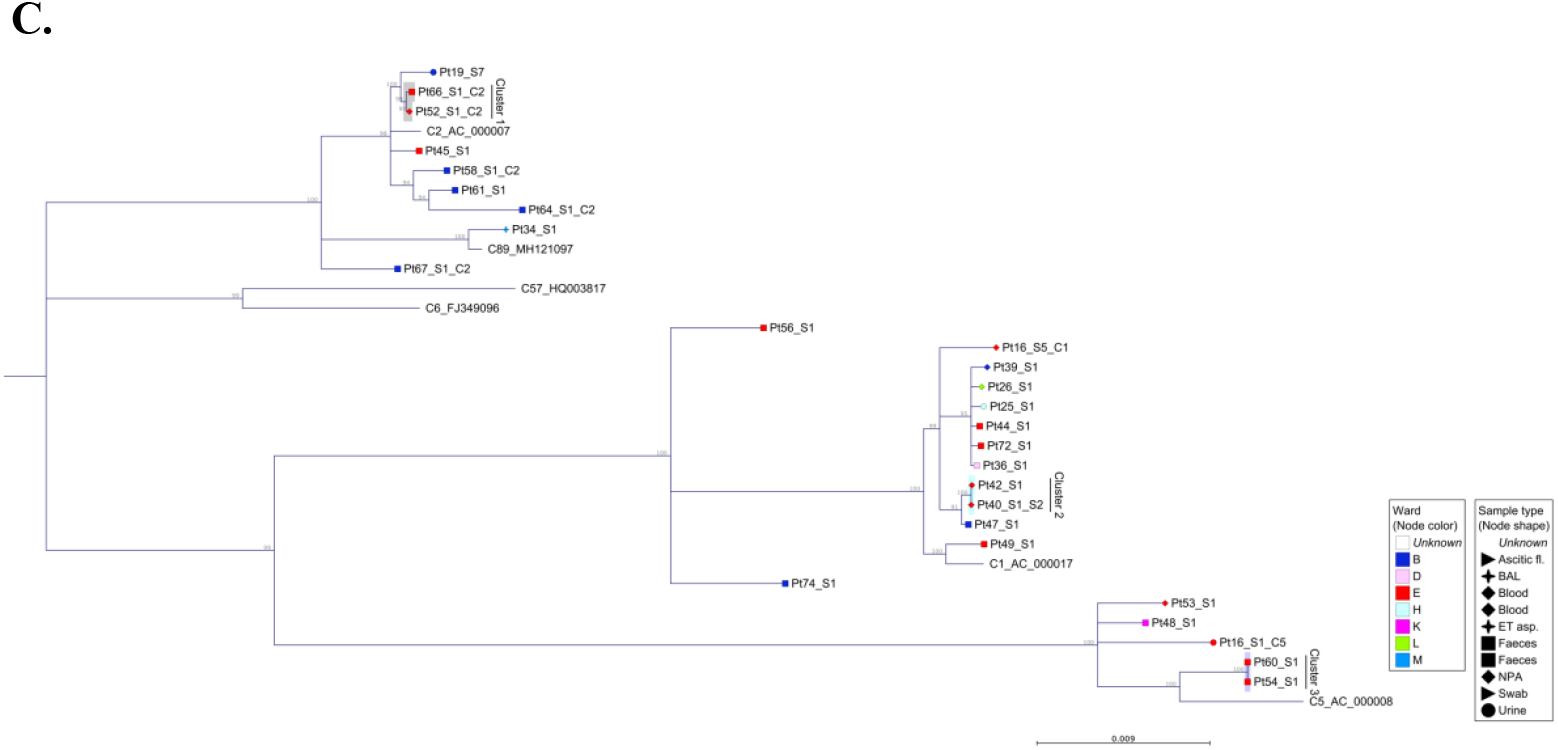
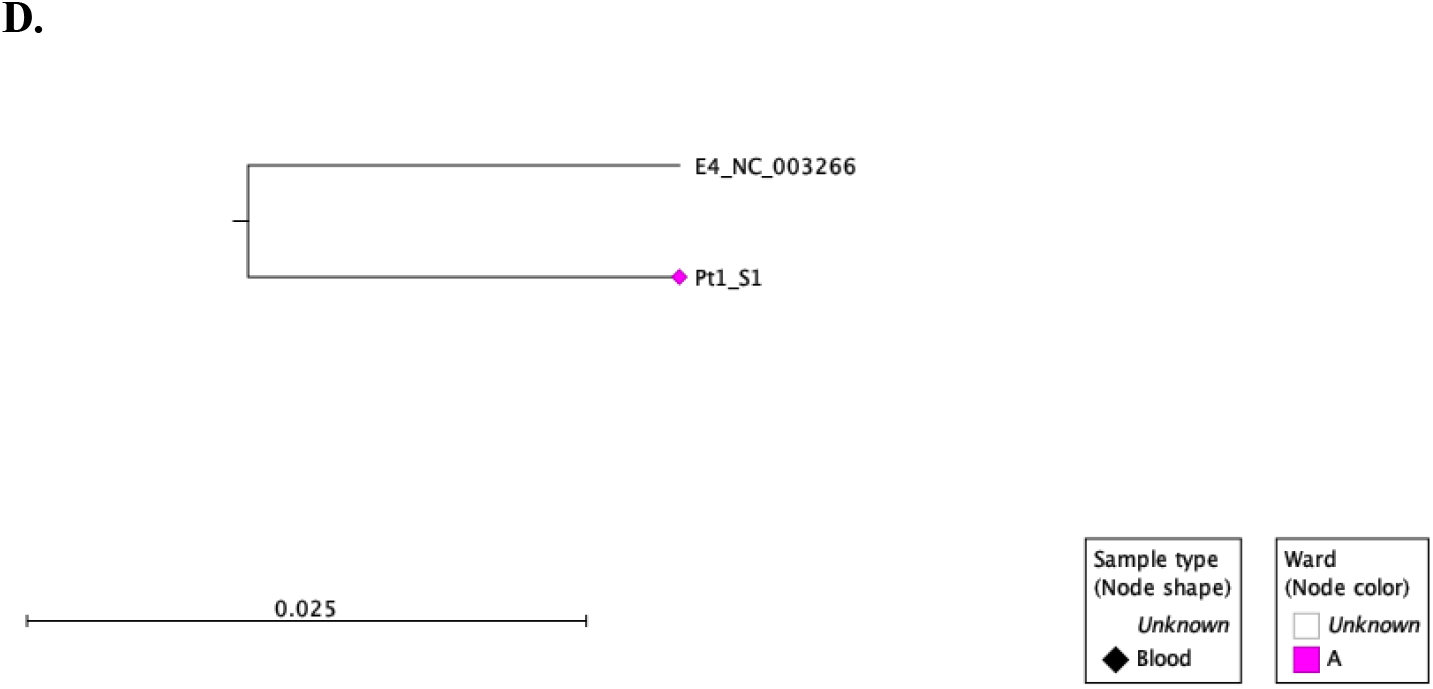
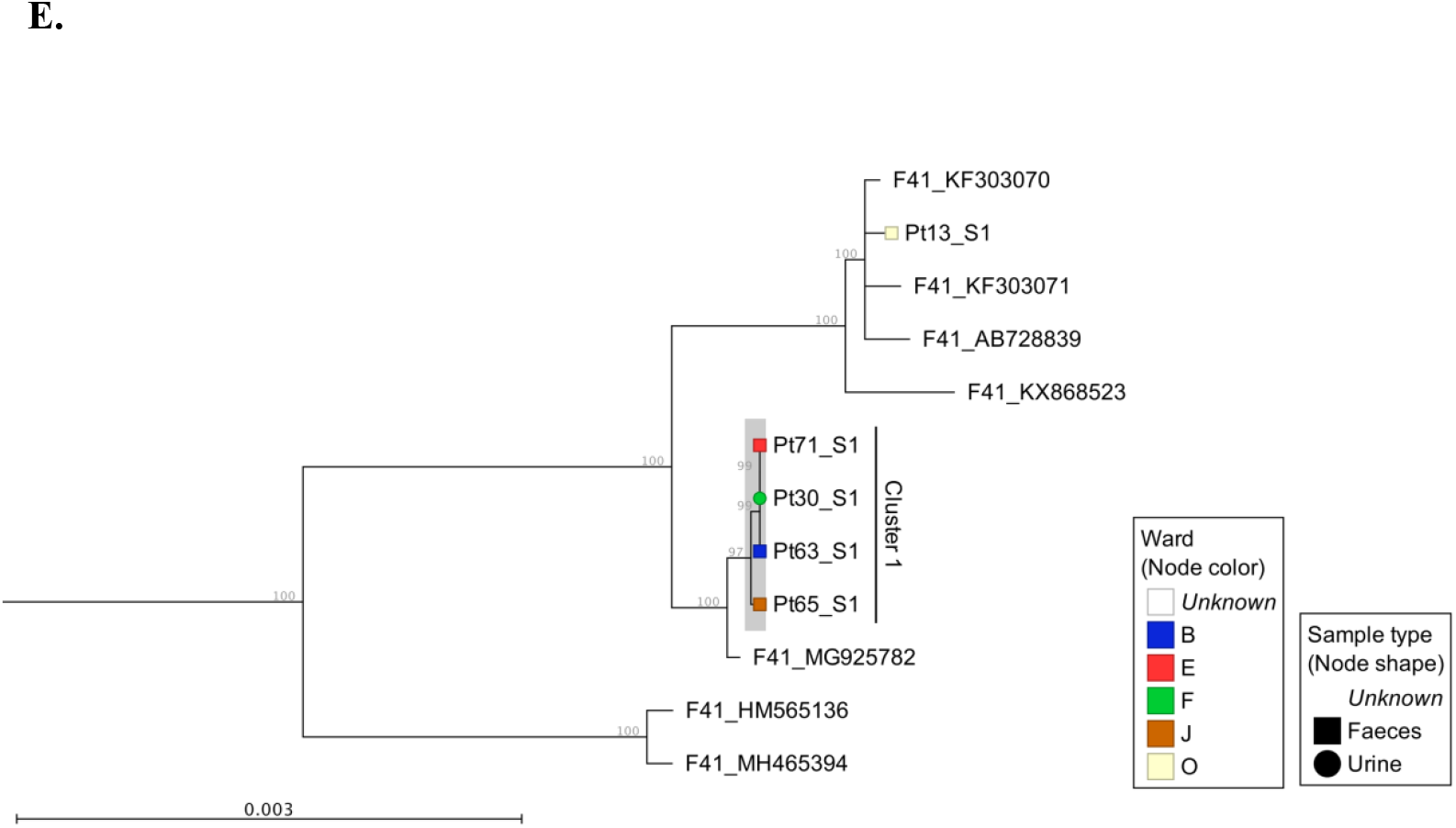
Maximum likelihood phylogenies of adenovirus full genome sequences, organised by species; (A.) species A viruses, (B.) species B viruses, (C.) species C viruses, (D.) species E and (E.) species F viruses. Sequences were aligned and maximum likelihood phylogenies generated using CLC Genomics Workbench (version 12.0.1), 500 bootstraps. Clinical samples are labelled according to their anonymised patient number (PtX) and specimen number (_SX). Additional samples from sequentially sampled patients have been collapsed. Inter-species reference sequences (A12, A18 and A61) were also removed from phylogeny A and from phylogeny I (F40) to aid visualisation of patient samples. The shape of each node correlates with the sample type and colour, the ward on which the sample was taken. Reference sequences are identified by their HAdV genotype and GenBank Accession. A bootstrap threshold of 80% is shown. Any clinical samples from at least two patients, with a bootstrap support greater than 90 were considered a cluster.

**Table 1.** Summary of Monophyletic Clusters Identified by Maximum Likelihood Phylogeny Using Whole HAdV Genome Sequences.

| HAdV type,<br>Sequence Cluster<br>Number | Sample Code | ICC<br>Number | IPC record | Ward Involved | Temporally<br>related<sup>a</sup> | Diversity within<br>cluster<sup>b</sup> | Conclusion |
| --- | --- | --- | --- | --- | --- | --- | --- |
| A31 Cluster 1 | Pt69_S1_A31 | - | HCAI not linked to outbreak | B | Yes | 0 | Confirmed transmission cluster |
| A31 Cluster 1 | Pt70_S1 | - | HCAI not linked to outbreak | E | Yes | 0 | Confirmed transmission cluster |
| A31 Cluster 1 | Pt68_S1 | 1 | Chronic HAdV – ICC 1 investigated | E | Yes | 0 | Confirmed transmission cluster |
| A31 Cluster 1 | Pt62_S1 | 1 | HCAI - ICC 1 investigated | B | Yes | 0 | Confirmed transmission cluster |
| A31 Cluster 1 | Pt57_S1 | 1 | HCAI - ICC 1 investigated | B | Yes | 0 | Confirmed transmission cluster |
| A31 Cluster 1 | Pt55_S1 | 1 | HCAI - ICC 1 investigated | E | Yes | 0 | Confirmed transmission cluster |
| A31 Cluster 2 | Pt11_S1 | - | Not classified | B | No | 6 | Likely transmission, unconfirmed |
| A31 Cluster 2 | Pt73_S1_A31 | - | HCAI | E | Yes | 3 | Confirmed transmission cluster |
| A31 Cluster 2 | Pt52_S1_A31* | - | HCAI | E | Yes | 3 | Confirmed transmission cluster |
| A31 Cluster 3 | Pt41_S1 | - | CAI | E | Yes | 1 | Confirmed transmission cluster |
| A31 Cluster 3 | Pt46_S1 | - | CAI | B | Yes | 1 | Confirmed transmission cluster |
| A31 Cluster 4 | Pt24_S1 | - | Not classified | E | Yes | 0-1 | Confirmed transmission cluster |
| A31 Cluster 4 | Pt8_S1 | - | Not classified | B | No | 0-1 | Confirmed transmission cluster |
| A31 Cluster 4 | Pt18_S1 | - | Probable HCAI | B | Yes | 0-1 | Confirmed transmission cluster |
| A31 Cluster 4 | Pt3_S1 | - | Not classified | B | Yes | 0-1 | Confirmed transmission cluster |
| B3 Cluster 1 | Pt27_S1 | - | Not classified | A | No | 13 | Unlikely transmission cluster |
| B3 Cluster 1 | Pt73_S1_B3 | - | Marked as long-term carriage from previous admission | E | No | 13 | Unlikely transmission cluster |
| C2 Cluster 1 | Pt52_S1_C2* | - | HCAI | E | No | 6 | Likely transmission, unconfirmed |
| C2 Cluster 1 | Pt66_S1_C2 | - | HCAI | E | No | 6 | Likely transmission, unconfirmed |
| C1 Cluster 2 | Pt42_S1 | - | CAI | E | Yes | 0 | Confirmed transmission cluster |
| C1 Cluster 2 | Pt40_S1_S2 | 2 | HCAI - ICC 2 investigated | E | Yes | 0 | Confirmed transmission cluster |
| C5 Cluster 3 | Pt60_S1* | - | Not classified | E | Yes | 0 | Confirmed transmission cluster |
| C5 Cluster 3 | Pt54_S1 | 1 | HCAI - ICC 1 investigated | E | Yes | 0 | Confirmed transmission cluster |
| F41 Cluster 1 | Pt30_S1 | - | Not classified | F | No | 0 | Confirmed transmission cluster |
| F41 Cluster 1 | Pt71_S1 | - | HCAI | E | Yes | 0 | Confirmed transmission cluster |
| F41 Cluster 1 | Pt63_S1 | - | HCAI | B | Yes | 0 | Confirmed transmission cluster |
| F41 Cluster 1 | Pt65_S1 | - | CAI | B | Yes | 3 | Likely transmission, unconfirmed |

Abbreviations: HAdV, human mastadenovirus; ICC, Infection Control Cluster; IPC, Infection, Prevention and Control; HCAI, healthcare-acquired infection; CAI, community acquired infection
<sup>a</sup>Temporally related, HAdV PCR positive whilst admitted to same/linked ward
<sup>b</sup>Diversity within cluster, expressed as the number of pairwise differences / single nucleotide polymorphisms (SNPs) across the whole genome
\*Indicates patients that died from or in association with overwhelming HAdV infection.

Four of the six patients from ICC one (patients 55, 57, 62 and 68) were phylogenetically linked (Figure 2A, A31 cluster one). An adsditional two patients (Pt69 and Pt70), documented as having HCAI, for whom no source of infection had previously been identified were linked phylogenetically to ICC one. One patient (Pt69) involved in monophyletic A31 cluster one had a mixed HAdV infection. The two patients from ICC two were not phylogenetically linked however, one of these patients, Pt40, was phylogenetically linked to another patient, Pt42, who had a concurrent HAdV-C2 infection. Whole genome sequencing identified an additional six monophyletic clusters, involving 17 patients that had not previously been identified by standard IPC follow-up (Figure 2 and Table 1).

Of the five patients who died (patients 52, 53, 59, 60 and 61) as a result of or in association with overwhelming HAdV infection (Supplementary Table 1), two were linked to a monophyletic cluster. Patient 52 was found to have a mixed genotype (C2 and A31) infection that was dominated by a phylogenetically unlinked C2 (Figure 2C) but with a minority subpopulation of A31 that clustered with two other patients (11 and 73, Figure 2A, A31 Cluster two). Patient 60 had a single C5 infection that clustered with patient 54 (C5 cluster three, Figure 2C). The remaining three patients had phylogenetically unlinked single genotype infections.

### Traditional Epidemiology, Contact Tracing Supported Phylogeny Assignments

Previous work has shown that infections can be linked over many years (23); A31 cluster four potentially transmitted over a five-year period (temporal relationship shown in Figure 3D). Using the new samples sequenced as part of this investigation, we confirmed the same, with a putative transmission cluster occurring over a three-year period (A31 cluster two, Figure 3B) and suggested prolonged transmission also occurring amongst other HAdV species; B3 cluster one over four years and F41 cluster one over three years (Figures 3E and 3I).

**Figure 3.**
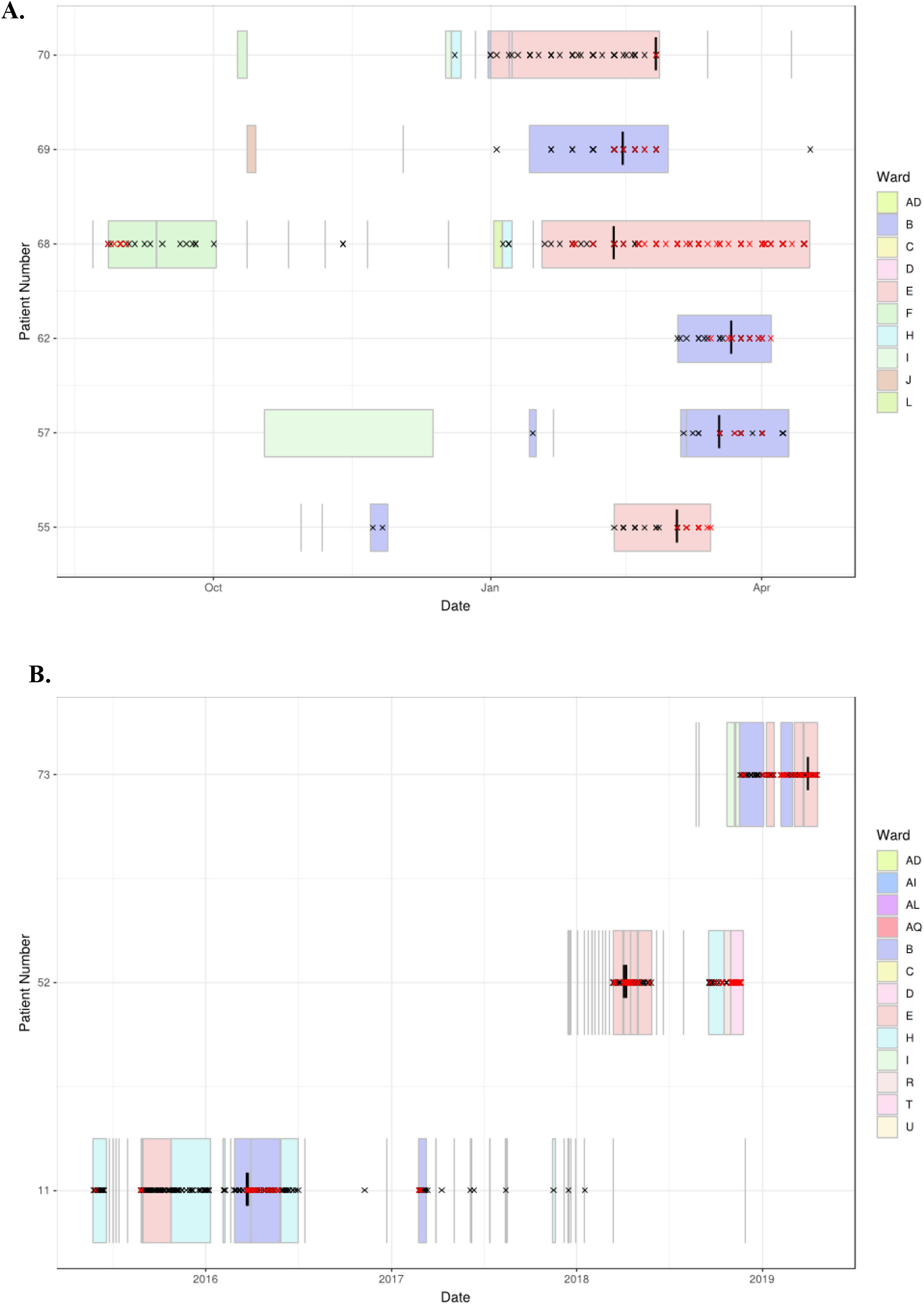
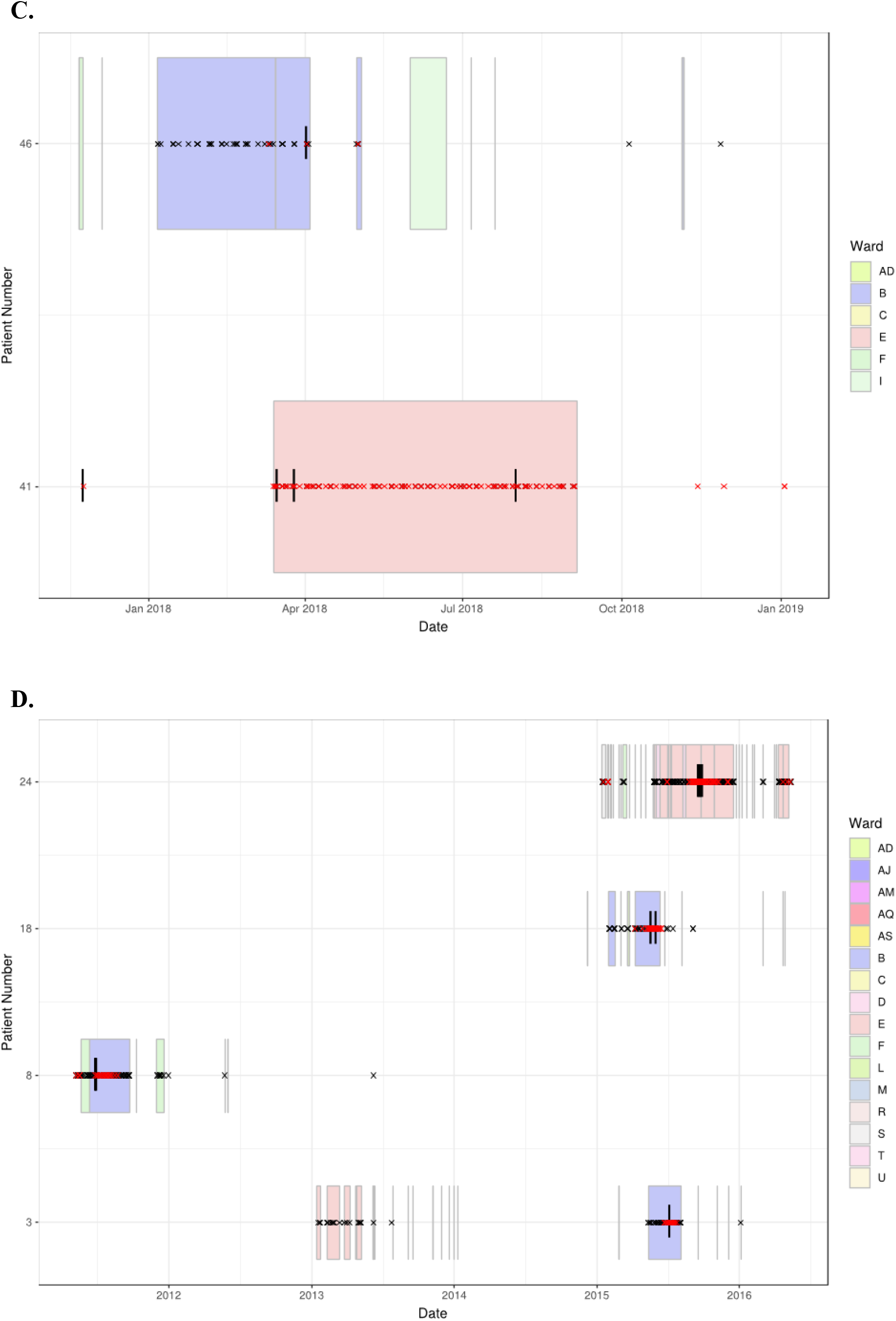
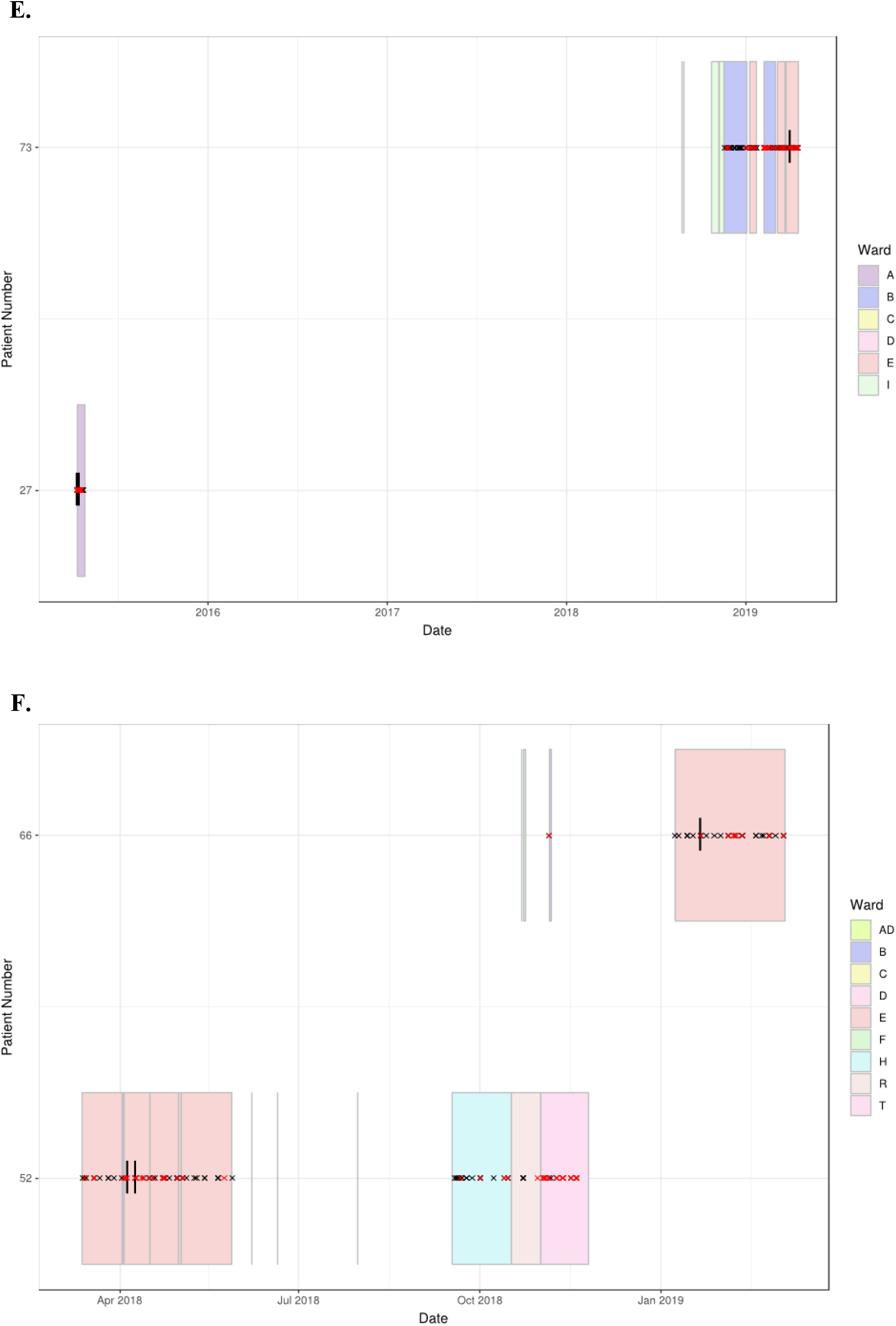
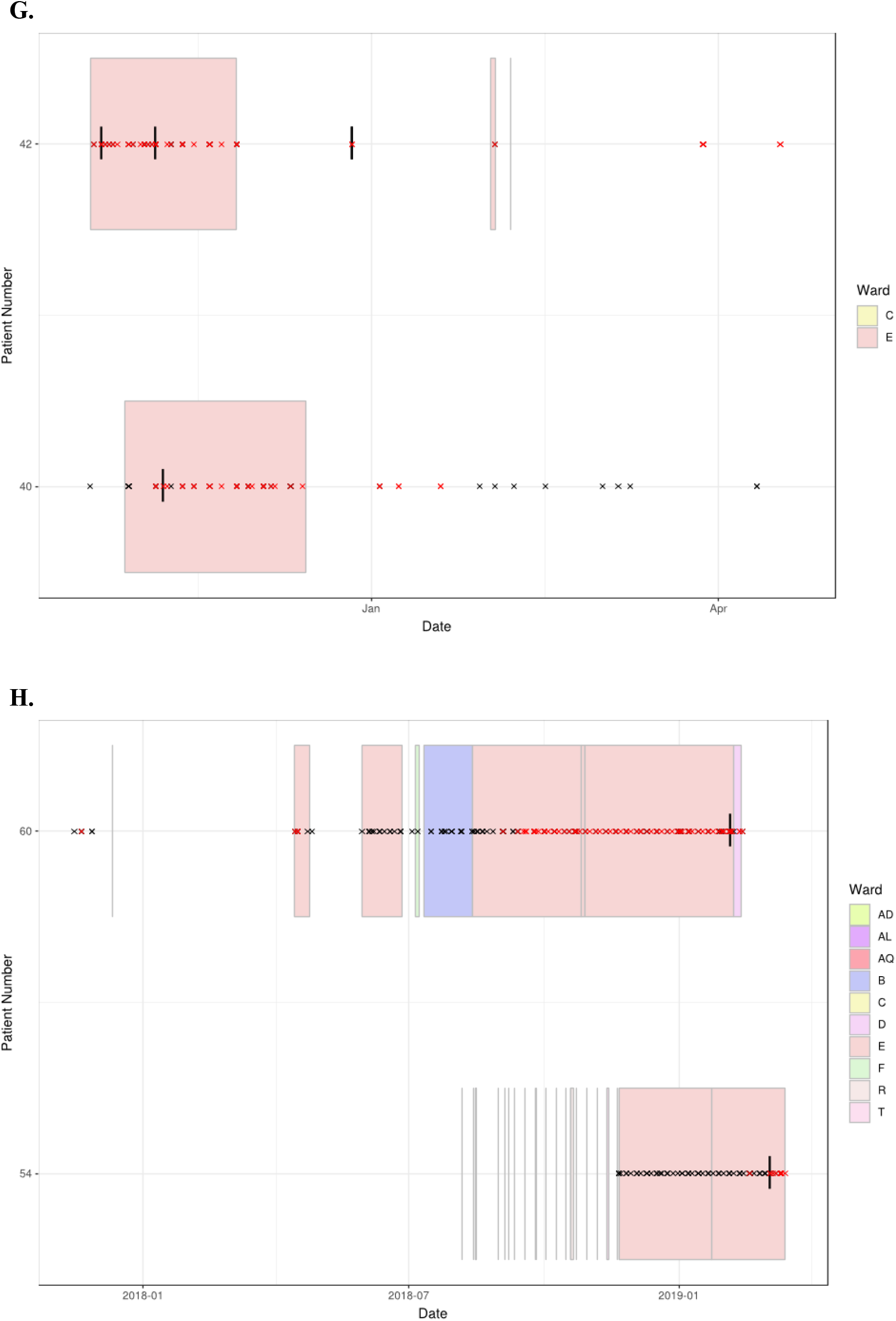
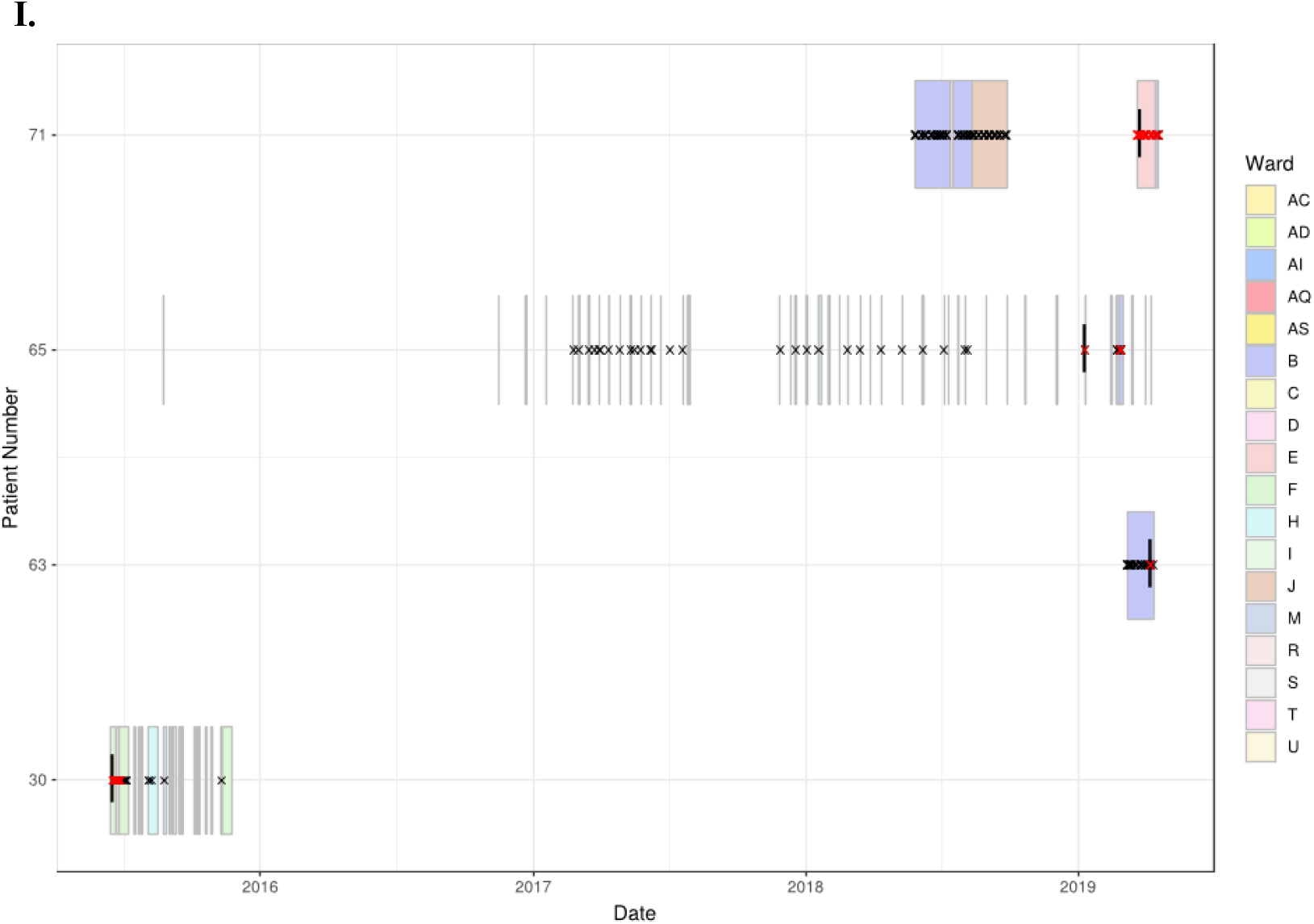
Temporal relationship between HAdV patients forming monophyletic clusters. Clinical data was extracted from hospital databases by the GOSH Digital Research Environment (DRE) team and linked to an anonymised patient number. The ggplot2 library was used to visualise data between positive adenovirus PCR results. Sequenced samples are indicated by a vertical black line, positive HAdV PCR samples by a red cross, negative HAdV PCR samples by a black cross and ward stays by coloured rectangles. (A) A31 cluster one, (B) A31 cluster two, (C) A31 cluster three, (D) A31 cluster four, (E) B3 cluster one, (F) C2 cluster one, (G) C1 cluster two, (H) C5 cluster three, (I) F41 cluster one.

Wards B and E were associated with all nine monophyletic clusters (Figures 2 and 3). As well as sharing clinical teams, these wards are joined by the same corridor and share facilities (dirty utility, kitchen, parents’ room and laundry), for this reason they are considered one HSCT unit. Four clusters containing ward B and E patients had temporal links with each other; A31 cluster one, A31 cluster three, C1 cluster two and C5 cluster three (Figure 3A, 3C, 3G, 3H and Table 1), supporting nosocomial transmission.

Three clusters contained one patient with no temporal links; patient 11 in A31 cluster two, patient 8 within A31 cluster four and patient 30 in F41 cluster one (Figures 3B, 3D and 3I). The two remaining clusters (C2 cluster one, Figure 3F and B3 cluster one, Figure 3E) did not share any temporal links with each other.

### Confidence in Genomic Links using Pairwise Distances

To quantify phylogenetic relationships, pairwise differences (single nucleotide polymorphisms (SNPs) between aligned consensus sequences) were calculated and grouped according to their epidemiological support (Figure 4). Epidemiologically linked monophyletic clusters were found to have ≤3 SNPs difference. This corroborated the number of differences previously defined for within host (≤2 SNPs, Figure 4C) and directly transmitted viruses (23), and further supports nosocomial transmission between clusters A31 cluster one, A31 cluster three, C1 cluster two and C5 cluster three (Table 1). Of the two patients (Pt54 and Pt60) involved in C5 cluster three, patient 60 died, however, this patient was admitted and HAdV PCR positive several months before patient 54 (Figure 3H) suggesting patient 60 may have been the source of this nosocomial infection.

**Figure 4.**
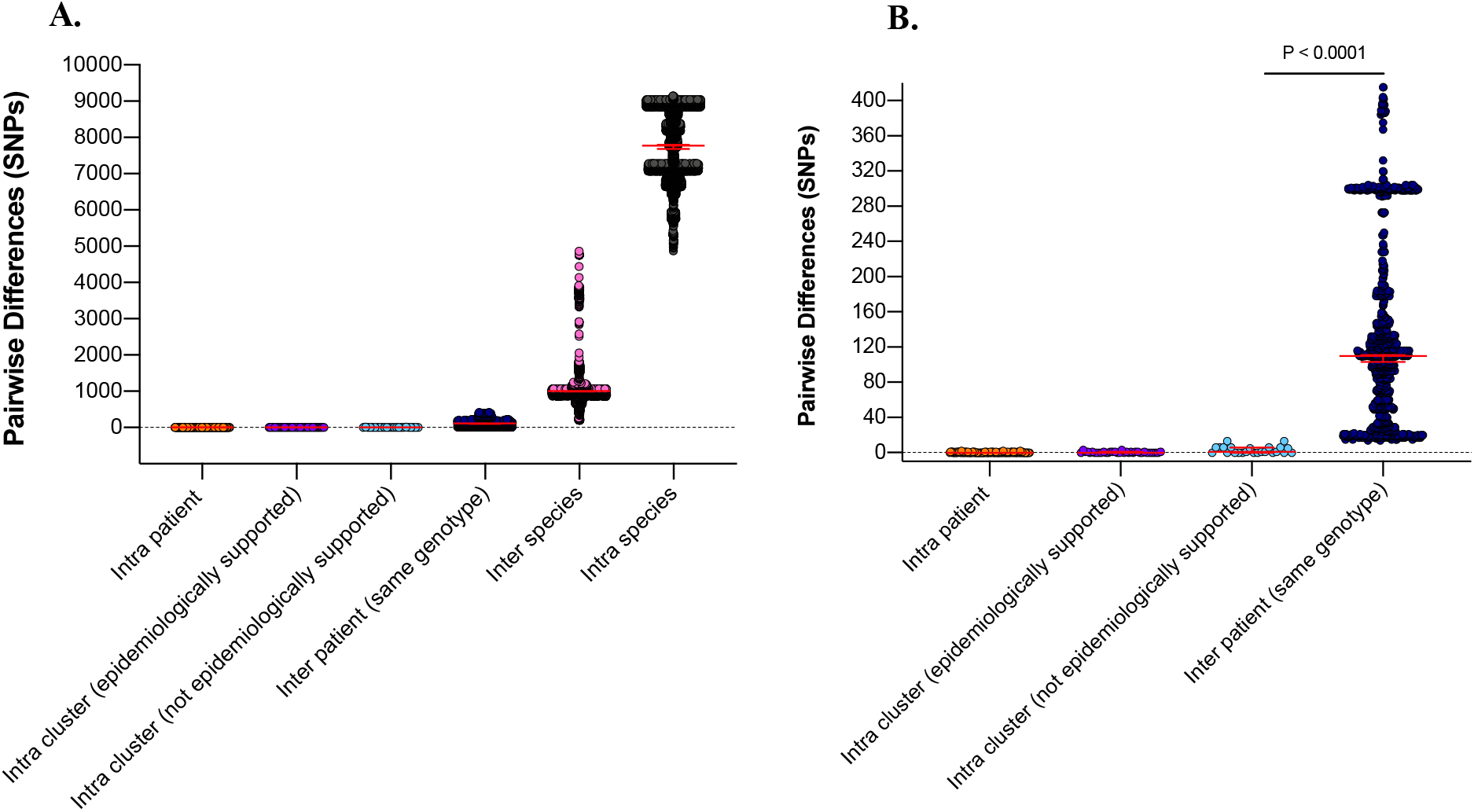
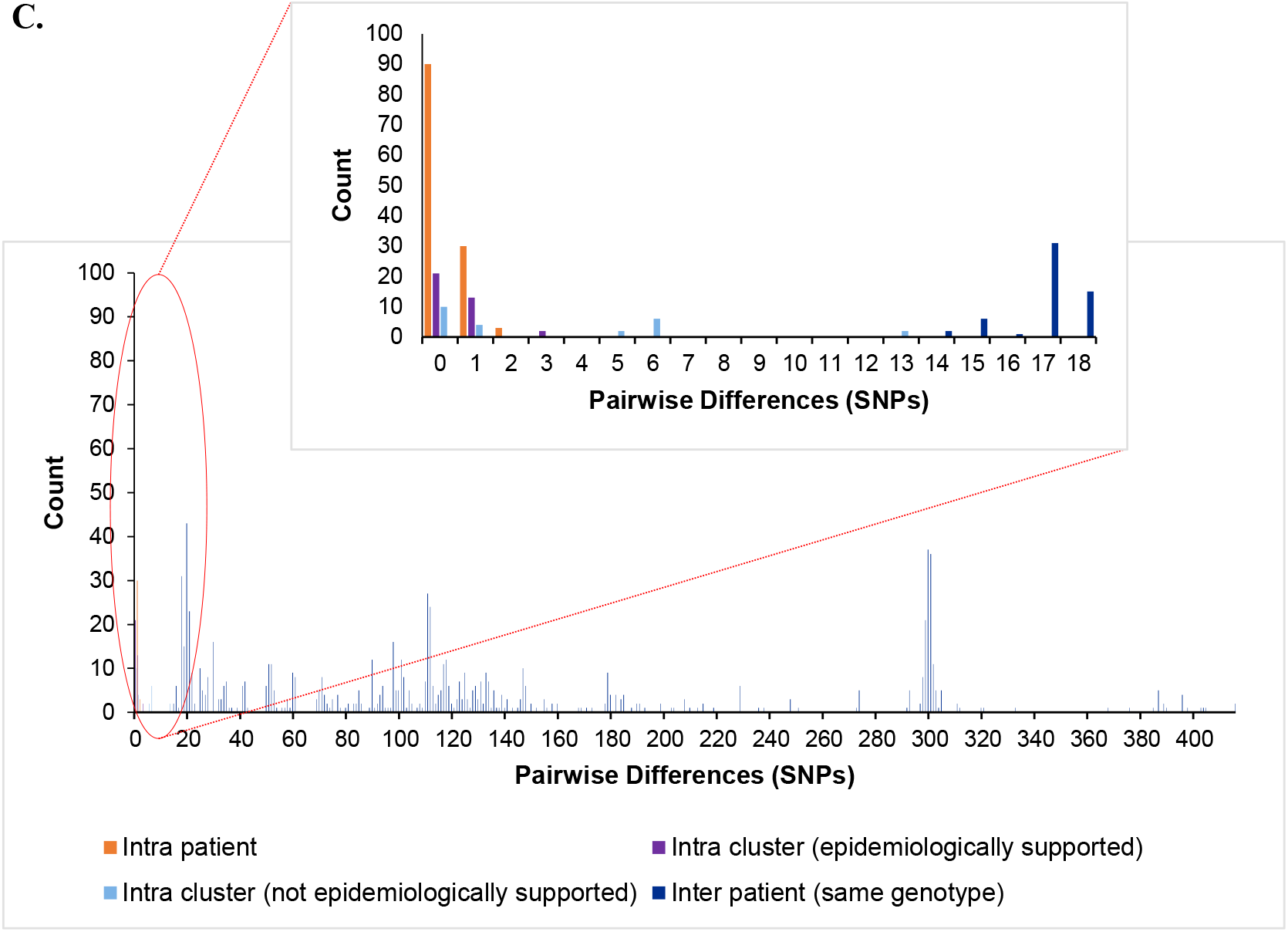
Pairwise differences equating to single nucleotide polymorphisms (SNPs) between clinical samples included in the study. (A.) Pairwise differences plotted by category; ‘within patient’, differences between samples taken from the same patient; ‘intra cluster (epidemiologically supported)’, differences between samples from different patients within a monophyletic cluster that are temporally linked (HAdV PCR positive whilst admitted to same/linked ward) by admission data; ‘intra cluster (not epidemiologically supported)’, differences between samples from different patients within a monophyletic cluster that are not linked temporally by admission data; ‘inter patient (same genotype)’, differences between samples from different patients within the same genotype; ‘inter species’, differences between samples from different patients within the same species but different genotypes and ‘intra species’, differences between samples from different patients between species. Median values with 95% confidence intervals are superimposed and plotted in red. (B.) Differences between the first four categories to aid visualisation. (C.) Highlighted precise count of pairwise differences found within first four categories.

Two monophyletic clusters, A31 cluster four and F41 cluster one, despite both containing one patient that was not linked temporally to the other patients (Figure 3D and 3I), differed by 0-3 SNPs suggesting nosocomial transmission (Table 1). With potentially unsampled patients or environmental intermediates, it is not possible to determine the route of transmission from these patients to the other patients within these clusters.

The remaining monophyletic clusters; A31 cluster two, B3 cluster one and C2 cluster one included at least one sequence separated by six to 13 SNPs. This range does not overlap with the number of SNPs found between unrelated patients of the same genotype (14-415 SNPs). However, interpretation of the epidemiological links between cases based on genomic data alone can be difficult because we do not currently understand the species-specific substitution rate of HAdV in chronically infected immunosuppressed patients.

B3 cluster one is the monophyletic cluster with the least support, without a temporal relationship between patients (admitted and tested four years apart) and 13 SNPs between the two sequences (14 SNPs were found between unrelated patient samples of the same genotype within species C viruses). Only three B3 infections were identified during the study period (92 and 93 SNPs separated this cluster from unrelated patient 14). The close clustering of these samples is therefore likely to be a result of the few publicly available UK HAdV B3 sequences. Further sequencing of HAdV-B3 genotypes is therefore required to substantiate the relationship found between these two patients.

## Discussion

This study demonstrates the major threat to immunosuppressed children that HAdV presents, providing a snapshot of a larger problem as only a minority of viruses were sequenced. With the level of infection present it is not possible to recognise HAdV outbreaks contemporaneously using conventional PCR methods. New infections identified by the diagnostic laboratory by PCR (Figure 1), are also likely to be over- or underrepresented, without genomic information it is impossible to know if these are genuine new infections or reactivation of clinically quiescent virus (30,31). Whole genome sequencing identified six patient clusters and at least 10 patient transmission events which were not identified using standard IPC investigations and whilst only focusing on high-risk patients from two wards. Rapid sequencing is now possible within 72 hours. This has already been shown to impact IPC management of SARS-CoV-2 in real-time (16) and would have impacted on the additional six patient clusters identified here.

The utility of WGS data is entirely dependent on the quality of sequences obtained. Poor genome coverage and low read depth generate tenuous links to other patients and any SNPs identified are poorly supported. Newly designed baits improved sequencing success (85% using version two baits, versus 42% using version one baits), especially for species C viruses [Supplementary Figure 8]. Species C HAdVs produce the most severe clinical manifestations amongst immunosuppressed patients, particularly those undergoing HSCT (32,33).

The high incidence of mixed infection (10% of patients) identified in this study highlights the superiority of WGS over PCR. Not only were patients incorrectly linked using standard methods, demonstrated by ICC two, HAdV PCR positive patients would go unnoticed if they later acquired a second HAdV co-infection. These secondary HAdV co-infections are important not only because of their role in transmission events; they could have different tissue tropisms, clinical consequence and provide a sentinel source of recombination (34–36).

This tertiary referral centre already has robust IPC precautions in place for high risk patients; single room isolation with en-suite facilities and environmental screening post discharge (22). Despite these precautions we were still able to confirm 21 patients (28%) were involved in a nosocomial transmission cluster, 15 patients (20%) ordinarily would have gone unnoticed or unlinked (Table 1.). Unidentified acquisition/transmission events can have clinically significant consequences including prolonged hospital stay, missed treatment opportunities and even death. Sequence data allowed us to investigate five HAdV associated deaths as part of the Trust patient safety review process. This is important for all cases of HCAI where the patient may have come to harm. The two HAdV associated deaths that were involved in transmission events here (Patient 52 and Patient 60) appeared to be the index case in their respective clusters (Figures 2A, 2C, 3B and 3H).

Urgent action needs to be taken to identify the source of HAdV acquisition in these patients in order to understand and halt transmission. Index cases may acquire their HAdV infection outside of hospital, but we have evidence that widely separated samples are linked; Pt8 within A31 cluster four, one SNP difference and Pt30 within F41 cluster one, zero SNPs (Figure 3D and 3I). In both cases the patients were immunocompromised and immunosuppressed long-term (X-linked lymphoproliferative disease and metastatic neuroblastoma) which is known to facilitate prolonged viral shedding (31). These patients, however, were discharged at least six months prior to related patients and sample positivity by PCR was absent for over three years. This suggests ongoing, undetected nosocomial transmission by unsampled intermediates which could include the environment, other patients, visiting relatives or staff members. Outbreaks amongst vulnerable patients already harbouring a high rate of HAdV positivity requires comprehensive surveillance. As a result, we have begun to implement routine HAdV WGS into a standard diagnostic algorithm to improve clinical care.

## Conclusions

The clinical utility of WGS technology for IPC purposes has begun to be realised for a number of important pathogens (12–16). Here we have demonstrated using a sensitive technique, that HCAI and mixed infection remains a significant problem despite the application of thorough IPC containment strategies. PCR alone fails to identify HAdV co-infection and transmission events, which can have catastrophic consequences amongst high risk patients. In order to combat this deficit, HAdV WGS is being implemented into routine diagnostics within this tertiary referral centre.

## Data Availability

Genome sequences will be assigned GenBank accessions upon publication.

## Acknowledgements

We acknowledge infrastructure support from the University College London Pathogen Genomics Unit, where the sequencing was conducted, and University College London Medical Research Council Centre for Molecular Medical Virology. Clinical samples were provided by Great Ormond Street Hospital for Children Departments of Microbiology, Virology and Infection Prevention and Control to the University College London Infection Bank. We are particularly grateful to John Booth, for his involvement in updating the DRE. The funders had no role in study design, data collection and interpretation, or the decision to submit the work for publication.

Potential conflict of interest: Agilent license our adenovirus bait designs.

